# Seek and You Will Find: A Prospective Case-Finding Study for the Detection of *Neisseria gonorrhoeae* Xpert CT/NG Diagnostic Escape Mutants in England

**DOI:** 10.64898/2026.11.11.26367844

**Authors:** Rachel Pitt-Kendall, Ella Finnie, Miriam O’Connor, Michael Rayment, Gary Whitlock, Marcus Pond, Malika Koulmi, Laura Viviani, Melissa Jansen van Rensburg, Katy Sinka, Derren Ready, Helen Fifer, Sarah Alexander

## Abstract

**Objectives:** In 2025, the UK Health Security Agency (UKHSA), confirmed the identification of a *Neisseria gonorrhoeae* (NG) diagnostic escape mutant for the Xpert® CT/NG assay (Cepheid). Subsequently, a small number of further mutants were reported internationally. The assay comprises two NG-specific targets, NG2 and NG4, and amplification is required in both to report a positive result. This prospective case-finding study aimed to investigate the presence of diagnostic escape mutants in NG-negative specimens from two sexual health services using the Xpert® CT/NG assay to provide in-clinic testing.

**Methods:** Residual Xpert® NG-negative clinical specimens were collected during December 2025 and transferred to the national STI Reference Laboratory at UKHSA. Following anonymisation extracts were tested on the GeneProof™ *Neisseria gonorrhoeae* PCR kit (GeneProof). GeneProof™ NG-positive specimens were re-tested on the Xpert® CT/NG assay to confirm.

**Results:** In total, 978 specimens were tested on the GeneProof™ assay; 99.7% (975/978) were confirmed as NG-negative. Three specimens (two rectal swabs and one urine sample) tested NG-positive on the GeneProof™ assay. Two specimens were re-tested on the Xpert® CT/NG assay, both re-tested NG-negative. Both samples amplified either the NG2 or NG4 target within the Xpert® NG assay, but not both targets required for a positive result. The remaining sample was insufficient for re-testing. An extrapolation of possible missed NG cases due to mutants, indicated that between 62 and 562 cases may have been missed in 2025 from these clinics.

**Conclusions:** This prospective case-finding study demonstrated excellent sensitivity of the Xpert® CT/NG assay with 99.7% of NG-negative samples correctly identified. However, a small number of diagnostic escape mutants were detected, and extrapolation suggests there may be some degree of missed cases from providers using this assay as a near-patient test. Further work is required to fully understand the prevalence of the mutants and clinical implications of missed diagnoses.

**Key messages:** *What is already known on this topic?:* *Neisseria gonorrhoeae* diagnostic escape mutants for the Xpert® CT/NG assay (Cepheid), lacking one or both diagnostic targets, have recently been identified in England and internationally. However, the true prevalence of these mutants and their impact on diagnoses are unknown.

*What this study adds:* Whilst 99.7% of specimens were confirmed as gonorrhoea-negative, this study identified two confirmed and one suspected diagnostic escape mutants among 978 Xpert® NG-negative specimens, from two sexual health services. These findings demonstrate that Xpert® NG escape mutants are present in clinic attendees in England, and may result in missed gonorrhoea diagnoses

*How this study might affect research, practice or policy:* The findings from this study support surveillance for diagnostic escape mutants and further studies to establish their prevalence and clinical impact.

## Introduction

Gonorrhoea, caused by *Neisseria gonorrhoeae* (NG), is the second most common sexually transmitted bacterial infection in England with 63,943 diagnoses reported in 2025, 57.6% of which were in gay, bisexual and other men who have sex with men (GBMSM) (1). Nucleic acid amplification tests (NAATs) offer superior sensitivity for diagnosis of infection to other methods, such as *in vitro* culture, particularly for extra-genital (2-6) and asymptomatic infections (4, 6, 7). Additionally, NAAT testing can use non-invasive, self-collected samples. Due to the potential for cross-reactivity of NAATs with commensal *Neisseria* species, especially in the pharynx, confirmation of positive results is recommended (2) using either a second target within an assay or a separate assay with a different target.

The Xpert® CT/NG assay (Cepheid) is a rapid (<2 h), modular cartridge-based NAAT that detects both *Chlamydia trachomatis* and NG DNA in one cartridge (4). The assay is frequently used as a confirmatory test as part of a two-step NG NAAT algorithm. The NG component of the assay has two chromosomal gonorrhoea-specific targets: NG2 and NG4, approximately 4 Kb apart, and amplification in both is required for the GeneXpert® (Cepheid) software to report a positive result. In June 2025, the national STI Reference Laboratory (STIRL) at the UK Health Security Agency confirmed the identification of a NG diagnostic escape mutant for the Xpert® CT/NG assay, following a likely recombination event with *Neisseria meningitidis* (8). Whole-genome sequencing revealed a complete absence of the NG2 and NG4 targets. A small number (n=16) of further mutants lacking one or both of the Xpert® NG targets were subsequently identified globally (8, 9). Due to the GeneXpert® software algorithm, single target mutants would also be reported as NG negative. Further analysis of these isolates indicated that recombination events had occurred in multiple distinct genomic lineages and was not the result of clonal spread (8, 9).

The true extent of use of the Cepheid GeneXpert® in England is unknown, however approximately 39.5% of participants of the UK NEQAS *C. trachomatis & N. gonorrhoeae* external quality assurance scheme reported use of this platform in 2026. Some sexual health services (SHSs), such as those delivered by the Chelsea and Westminster NHS Foundation Trust in London, offer in-clinic testing with the Xpert® CT/NG assay. These SHSs serve a substantial proportion of SHS attendees nationally and reported 13.4% of the total NG diagnoses from England in 2025 (8,556/63,943) (10).

We undertook an anonymised prospective case finding study of Xpert® NG-negative specimens from individuals attending two Chelsea and Westminster NHS Foundation Trust SHSs, to estimate the prevalence of these diagnostic escape mutants.

## Methods

### Sample size calculations

Assuming the prevalence of Xpert® NG diagnostic escape mutants was ≤0.3%, it was calculated that at least 998 NG-negative specimens would need to be tested to detect at least one mutant with 95% confidence.

### Specimen Collection period

Owing to differences in testing numbers, sampling periods were tailored to each SHS. Clinic 1 which performed an average of 3,842 NG tests per month in 2025, collected samples during the first week of December 2025 only. Clinic 2 which averaged 959 NG tests per month, collected samples throughout December 2025. To ensure representation of different specimen types, only one sample type (e.g. vaginal swab) was selected per day at Clinic 1 and per week at Clinic 2. Both clinics provide services to individuals of all sexes, gender identities and sexual orientations. However, Clinic 1 serves a higher proportion of GBMSM.

### Sample processing

Residual Xpert® NG-negative specimens (urine, genital or rectal swabs) in Xpert® CT/NG sample collection buffer (Cepheid), were forwarded to STIRL. Pharyngeal swabs were excluded due to known cross-reactivity of diagnostic assays with commensal *Neisseria* at this site. Samples from individuals attending for test-of-cure were also excluded.

Specimens were anonymised (retaining sample site and age), aliquoted and DNA extracted using the QIASymphony and DSP Virus/Pathogen Midi kit (Qiagen). Extracts were tested using the GeneProof™ *Neisseria gonorrhoeae* PCR kit (GeneProof) on the QuantStudio 5 (Applied Biosystems), as per manufacturer’s instructions. The assay targets both the NG *porA* pseudogene and 16S rRNA gene, reporting in the same channel to provide increased sensitivity of detection in specimens with low DNA load.

GeneProof™ NG-positive specimens were retested following re-extraction where possible, or on the original extract if insufficient volume remained. The limit of detection of the GeneProof™ assay was established locally as 30 colony forming units/mL, equating to a cycle threshold (Ct) value of 37. As such results were interpreted as follows:

- Positive/Positive (at least one Ct: ≤37) – NG-positive
- Positive/Positive (both Ct: >37) – NG-indeterminate
- Positive (any Ct)/Negative – NG-indeterminate
- Negative – NG-negative

Specimens classified as NG-positive were re-tested on the Xpert® CT/NG assay on the GeneXpert® platform. No further testing was performed for NG-indeterminate specimens due to expected low DNA concentration. Given the inconsistent amplification of NG-indeterminate samples, NG-negative and NG-indeterminate specimens were grouped together for the overall analysis to ensure a conservative interpretation of the findings.

### Estimated annual false-negative Xpert® NG reports

To estimate the potential number of gonorrhoea infections that may have been missed in 2025 due to Xpert® NG escape mutants, the prevalence of confirmed and suspected mutants identified in this study was applied to the total number of NG tests performed at the participating SHSs. Suspected mutants were GeneProof™-positive samples that could not undergo repeat testing with the Xpert® CT/NG assay. The numbers of NG tests performed, and NG diagnoses reported in 2025 by the two SHSs were extracted from the GUMCAD STI Surveillance System (10). The upper and lower 95% confidence interval limits were used to estimate a plausible range of missed infections.

### Ethical approval

This study was undertaken for health protection purposes. This study did not involve human participants. All clinical samples tested were residual, anonymised, and unlinked from any identifiable information prior to testing.

## Results

### Sample processing

In total 978 Xpert® NG-negative clinical specimens (179 vaginal swabs, 397 rectal swabs and 402 urine specimens) were tested on the GeneProof™ *Neisseria gonorrhoeae* PCR kit. The majority of which (975/978, 99.7%) were confirmed as NG-negative or returned an NG-indeterminate result on the alternate assay (Table 1). Three specimens (3/978, 0.3%), two rectal swabs and one urine sample, from three individuals (inferred from patient age) from the same clinic, tested positive on the GeneProof™ assay (Table 1).

**Table 1.** Summary of the NG-negative specimens received, GeneProof™ *Neisseria gonorrhoeae* PCR kit (GeneProof) results and Xpert® CT/NG (Cepheid) re-test results

|  | GeneProof<sup>TM</sup> *Neisseria gonorrhoeae* PCR |  |  |  | Xpert® CT/NG assay re-test |  |  |
|---|---|---|---|---|---|---|---|
|  | NG-negative* |  | NG-positive |  | NG-negative (n=2) |  | NG-positive |
|  | N | % | N | % | NG2+/NG4- | NG2-/NG4+ | N |
| Vaginal (n=179, 18.3%) | 179 | 100 (95% CI: 97.9-100) | 0 | 0 |  |  |  |
| Rectal (n=397, 38.8%) | 395 | 99.5 (95% CI: 98.2-99.9) | **2** | 0.5 (95% CI: 0.1-1.8) | 1 | 1 | 0 |
| Urine (n=402, 41.1%) | 401 | 99.8 (95% CI: 98.6-100) | **1\*\*** | 0.3 (95% CI: 0.04-1.4) | n/a** |  |  |
| Total (n=978) | 975 | 99.7(95% CI: 99.1-99.9) | 3 | 0.3 (95% CI: 0.1-0.9) |  |  |  |

\*including NG-indeterminate specimens; \*\*insufficient sample volume remained for re-testing on the Xpert® CT/NG assay (Cepheid)

### Xpert® CT/NG assay re-testing

Sufficient residual volume remained for two of the NG-positive samples (both rectal) to allow re-testing on the Xpert® CT/NG assay. Both samples retested NG-negative (Table 1). Review of the individual target amplification reports revealed that either NG2 or NG4 was amplified (Table 1), but not both, which is required for a positive Xpert® result. Therefore at least 2/978 (0.2%) of the specimens analysed were confirmed as false-negative results on the Xpert® CT/NG assay, and had result profiles consistent with previously detected diagnostic escape mutants (8, 9). The remaining specimen that was insufficient for re-testing was classified as a “suspected” mutant.

### Estimated annual false-negative Xpert® NG reports

In 2025, the two sexual health clinics participating in this study performed 62,404 NG tests, of which 7,396 (11.9%) were reported as NG-positive (10). Using the lower and upper 95% confidence limits reported in Table 1, it was estimated that between 62 and 562 tests were falsely reported as NG-negative due to the presence of an escape mutant in 2025.

## Discussion

This prospective case-finding study investigated NG diagnostic escape mutants for the Xpert® CT/NG assay in two large SHSs using the assay in clinic as a near-patient diagnostic test. Re-testing of NG-negative specimens with an alternate assay at the national STI Reference Laboratory confirmed the original NG-negative result for 99.7% of specimens (including specimens categorised as NG-indeterminate). However, two (0.2%) single-target Xpert® NG mutants containing specimens were detected. A further specimen was classified as a possible mutant. All confirmed and possible NG-mutants were sourced from different patients attending the same clinic (Clinic 1). In this analysis, NG-indeterminate specimens were grouped with the NG-negative samples as evidence of NG-specific DNA could not be reliably detected in these specimens using the GeneProof™ assay. This may be due to differences in the analytical sensitivity of the tests or cross-reactivity of the GeneProof™ assay as well as false-negative results on the Xpert® NG assay.

The requirement for both NG assay targets (NG2 and NG4) to detect amplification to return a positive result is assumed to enhance specificity and prevent over-reporting where amplification in one target is due to cross-reactivity with other *Neisseria* species, a well-known problem in NG diagnostics. However, this cautious approach results in vulnerability to under-reporting of true NG diagnoses, where a single-target mutant is present, such as those detected in this study. The GeneProof™ assay contains two NG-specific targets that are different to the targets within the Xpert® NG assay. However, both report in the same channel therefore it is not possible to discern whether one or both targets have amplified. For the two confirmed mutant specimens, on review of the individual Xpert® sample reports, a positive signal was recorded for either the NG2 or NG4 target. Therefore, the samples were positive for at least two independent NG-specific targets, indicating that the NG-positive results were genuine, and not a result of cross-reactivity.

Although this study was not designed to determine the prevalence of Xpert® CT/NG escape mutants, our findings suggest that some under-reporting of gonorrhoea diagnoses may occur in clinics using the Xpert® CT/NG assay. Reassuringly, the vast majority of samples were confirmed as NG-negative. Further studies involving a broader and more geographically diverse range of clinics are required to better understand the prevalence and distribution of these mutants.

The samples included in this study were anonymised prior to testing. Whilst this was a pragmatic approach as testing was not performed in real-time, it did limit the interpretation of the findings as it was not possible to review the patient records to understand if NG had been diagnosed through other means, and assess the potential impact of the false-negative report. Symptomatic patients would be subject to clinical follow-up including repeat testing. Therefore, it is likely that infections within this group would be captured in real-time and treated appropriately. A further limitation of this study was exclusion of *Chlamydia trachomatis*-positive, NG-negative rectal swabs which were unavailable due to referral to a separate laboratory for detection of Lymphogranuloma venereum genovars (as per current national guidance (11)). Future studies should incorporate these important samples so as not to restrict prevalence estimates. Finally, this study utilised residual clinical specimens, which whilst convenient, prevented further characterisation of the mutants detected due to lack of sample volume for additional testing. Future studies could consider testing cultured isolates in place of clinical specimens to enable genome sequencing and other downstream characterisation of putative escape mutants.

Overall, the study demonstrates the high sensitivity of the Xpert® CT/NG assay. A small number of diagnostic escape mutants were detected and extrapolations from this indicate that there may be some missed gonorrhoea diagnoses from providers using this assay as a primary diagnostic test. Further work is required to comprehensively investigate the true prevalence of these mutants, and any clinical impact of under-reporting.

## Data Availability

All data produced in the present work are contained in the manuscript

## Acknowledgements

The authors would like to acknowledge Suzy Sun for assistance with GUMCAD data and Jack Minshull for help with the co-ordination of laboratory testing.

## Contributors

RPK, SA, JS, MR, GW and MJvR conceived the study protocol. EF, MK, RPK and MOC performed laboratory testing and data analysis. RPK, EF and SA prepared the original manuscript. All authors reviewed, edited and approved the final manuscript.

## References

1. UK Health Security Agency. Table 4 All STI diagnoses and services by gender and sexual orientation. Sexually transmitted infections (STIs): annual data. https://www.gov.uk/government/statistics/sexually-transmitted-infections-stis-annual-data-tables2025.

2. Fifer H, Ismail MA, Soni S, Nwaosu U, Sadiq ST, Milligan A, et al. British Association of Sexual Health and HIV UK National Guideline for the Management of infection with Neisseria gonorrhoeae, 2025. Int J STD AIDS. 2025:9564624251345195.

3. Cornelisse VJ, Chow EP, Huffam S, Fairley CK, Bissessor M, De Petra V, et al. Increased Detection of Pharyngeal and Rectal Gonorrhea in Men Who Have Sex With Men After Transition From Culture To Nucleic Acid Amplification Testing. Sex Transm Dis. 2017;44(2):114–7.

4. Gaydos CA, Van Der Pol B, Jett-Goheen M, Barnes M, Quinn N, Clark C, et al. Performance of the Cepheid CT/NG Xpert Rapid PCR Test for Detection of Chlamydia trachomatis and Neisseria gonorrhoeae. J Clin Microbiol. 2013;51(6):1666–72.

5. Levy V, Blackmore CS, Klausner JD. Self-collection of specimens for nucleic acid-based diagnosis of pharyngeal, cervicovaginal, urethral, and rectal Neisseria gonorrhoeae and Chlamydia trachomatis infections. Methods Mol Biol. 2012;903:407–18.

6. Geelen TH, Rossen JW, Beerens AM, Poort L, Morre SA, Ritmeester WS, et al. Performance of cobas(R) 4800 and m2000 real-time assays for detection of Chlamydia trachomatis and Neisseria gonorrhoeae in rectal and self-collected vaginal specimen. Diagn Microbiol Infect Dis. 2013;77(2):101–5.

7. Palmer HM, Mallinson H, Wood RL, Herring AJ. Evaluation of the specificities of five DNA amplification methods for the detection of Neisseria gonorrhoeae. J Clin Microbiol. 2003;41(2):835–7.

8. Jansen van Rensburg M, Pitt-Kendall R, Hincke M, Cliff PR, Shaw J, Sinka K, et al. Detection and characterisation of Xpert CT/NG assay Neisseria gonorrhoeae diagnostic escape mutants, England, June 2025. Euro Surveill. 2025;30(36).

9. Golparian D, Bazzo ML, Gaspar PC, Unemo M. Global distribution of Xpert CT/NG assay Neisseria gonorrhoeae escape variants indicates sporadic emergence with limited clonality between 2016 and 2025. Euro Surveill. 2025;30(46).

10. UK Health Security Agency. GUMCAD STI Surveillance System. https://www.gov.uk/guidance/gumcad-sti-surveillance-system.; 2026.

11. White J, O’Farrell N, Daniels D, British Association for Sexual H, Hiv. 2013 UK National Guideline for the management of lymphogranuloma venereum: Clinical Effectiveness Group of the British Association for Sexual Health and HIV (CEG/BASHH) Guideline development group. Int J STD AIDS. 2013;24(8):593–601.

